# Clinical Utility of Combined Optical Genome Mapping and 523-gene Next Generation Sequencing Panel For Comprehensive Evaluation of Myeloid Cancers

**DOI:** 10.1101/2022.01.15.22269355

**Authors:** Nikhil Shri Sahajpal, Ashis K Mondal, Sudha Ananth, Daniel Saul, Soheil Shams, Alex R Hastie, Natasha M. Savage, Vamsi Kota, Alka Chaubey, Ravindra Kolhe

## Abstract

The standard-of-care (SOC) for genomic testing of myeloid cancers primarily relies on karyotyping and fluorescent in situ hydridization (FISH) (cytogenetic analysis) and targeted gene panels (≤54 genes) that harbor hotspot pathogenic variants (molecular genetic analysis). Both cytogenetic and molecular testing workup is necessary for the identification and detection of large structural variants (SVs) and small variants like single nucleotide variants (SNV) and indels, respectively. Despite this combinatorial approach, ∼50% of myeloid cancer genomes remain cytogenetically normal, and the limited sequencing variant profiles obtained from targeted panels are unable to resolve the genetic etiology of these myeloid tumors. In this study, we evaluated the performance and clinical utility of optical genome mapping (OGM) and a 523-gene next-generation sequencing (NGS) panel for comprehensive genomic profiling of 15 myeloid tumors and compared it to SOC cytogenetic methods (karyotyping and FISH) and a 54-gene NGS panel. OGM and the 523-gene NGS panel were found to have an analytical concordance of 100% with karyotyping, FISH, and the 54-gene panel, respectively. Additionally, OGM better characterized and resolved the structural variants previously reported by karyotyping in five cases, such as identifying the genomic content of marker and ring chromosomes. OGM also identified several additional translocations and eleven copy number variations (CNVs), of which the CNVs were validated/confirmed by the 523-gene panel. The 523-gene panel identified seven additional clinically relevant SNVs (two tier 1A variants and five tier 2C variants, as per the ACMG/AMP guidelines) in four cases. The simultaneous visualization of SVs and small NGS detected sequence variants (SNVs and small indels) from OGM and 523-gene NGS panel, respectively in the NxClinical software v6.1 identified two clinically relevant compound heterozygous events in two samples. This study demonstrates the higher sensitivity, resolution, accuracy, and ability to reveal cryptic and clinically relevant novel variants in myeloid cancers as compared to SOC methodologies. Our cost-effective approach of using OGM and a 523-gene NGS panel for comprehensive genomic profiling of myeloid cancers will not only increase the yield of actionable targets leading to improved clinical outcomes but also help resolve our ongoing conundrum of apparently genomically normal myeloid cancers by providing more answers.

## Introduction

Myeloid malignancies are characterized by uncontrolled proliferation and/or defects in the differentiation of abnormal myeloid progenitor cells. Myelodysplastic syndromes (MDS) and myeloproliferative neoplasms (MPN) are often thought to be precursors to higher-grade myeloid malignancies, i.e. acute myeloid leukemia (AML) [**1**]. The National Comprehensive Cancer Network (NCCN) guidelines for the genetic diagnosis of MDS/MPN/AML recommends bone marrow cytogenetic analysis using karyotyping and fluorescence in situ hybridization (FISH), in addition to molecular analysis for at least *JAK2, CALR*, and *MPL* genes for MPN and *c-KIT, FLT3* (ITD and TKD), *NPM1, CEBPA* (biallelic), *IDH1*, and *IDH2* genes for AML. The guidelines further recommend molecular testing using multiplex gene panels and next-generation sequencing (NGS) analysis for comprehensive prognostic assessment [**2**].

In this context, the routine assessment of myeloid cancers in current clinical care incorporates karyotyping and FISH [occasionally chromosomal microarrays (CMA)] for cytogenetic analysis [**2-4**], and targeted gene panels (≤54 genes) that screen prominent hotspot pathogenic variants for molecular characterization [**5**,**6**]. This approach has been widely implemented but suffers from several limitations that include: first, a combination of traditional cytogenetic methods (karyotype and FISH/CMA) are required to obtain structural variation analysis according to the guidelines, as each technology has limitations. Karyotyping is sufficient for genome-wide structural variation (SV) detection but has low-resolution with a maximum banding resolution of ∼5-10 Mb. FISH provides higher resolution but is targeted and not appropriate for genome-wide analysis. CMA has the highest sensitivity to detect copy number variations (CNV) but cannot detect balanced SV, or determine the location or orientation of copy number gain (or amplification) regions of the genome. Second, the use of small-targeted gene panels for molecular characterization yields an incomplete sequence variant profile (omitting several important tumorigenic sequence variants) [**7**]. Despite this standard of care workflow including both cytogenetic and molecular methods, approximately 50% of the myeloid cancers remain cytogenetically normal [**8-10**] and insufficient sequencing variants profile are unable to resolve the heterogeneity in diagnostic features and/or outcomes in patients with myeloid cancers [**5**,**6**,**7**,**11**]. In summary, we have been struggling with severe limitations of our current testing modalities for adequate evaluation of myeloid malignancies. On multiple occasions, with our existing testing methods (karyoptying/FISH/CMA/54-NGS) patients with myeloid malignancies harboring up to 80-90% blasts end up with normal genome reports. This has consistently led to suboptimal diagnostic or prognostic classification and inadequate therapy/transplant selection, which ultimately leads to poor clinical outcomes.

Recently, in the cytogenetic domain, optical genome mapping (OGM) has emerged as a next-generation cytogenomic technology that can detect all classes of SVs (insertions, deletions, duplications, inversions, translocations), and complex rearrangements at a higher resolution than the standard of care methods [**12-17**]. The OGM technique is based on imaging ultra-long DNA (>150 kbp) molecules labelled at specific sequence motifs (CTTAAG) that span the entire genome (on average, every 5 kbp). Generation of OGM data at approximately 400x coverage allows for the detection of low-level SVs as required for many meyelogenous malignancy samples since cancer cells will be analyzed through a background of stromal cells in blood and bone marrow. Recently, we along with other groups evaluated OGM’s potential and role for the detection of SVs in patients with hematological malignancies and found 100% clinical concordance with traditional cytogenetic analysis [**15-17**]. In addition, OGM was able to provide higher resolution and resolve or refine previously identified aberrations and identified additional clinically relevant abnormalities that remained beyond the purview of current methods [**15-17**].

In the molecular domain, targeted NGS panels (≤54 genes) are primarily employed as they generate high coverage (∼500x) and are cost and time-effective with minimal data analysis and reporting complexity. Recently, whole-genome sequencing (GS) at ∼50-120x read depth analysis has been reported to identify targeted SVs, karyotype level CNVs, and sequencing variants in myeloid neoplasms [**18**]. However, the authors reported GS to be time and cost-prohibitive requiring sophisticated instrumentation and bioinformatics (data analysis, interpretation, and reporting) challenges, and also reported false-negative sequence variants compared to the 40-gene NGS panel [**18**]. For widespread utilization, a sequencing strategy should provide a thorough molecular characterization with minimal data analysis and enable easy reporting. In this regard, we have previously validated a 523-gene NGS panel for simultaneous analysis of sequence variants, tumor mutation burden (TMB), and microsatellite instability (MSI) for myeloid cancers [**7**].

We hypothesize that a combination of OGM and a 523-gene NGS panel would provide a novel and superior approach for the comprehensive characterization of cytogenetic and molecular variants in myeloid malignancies. In this proof-of-principle study, we evaluated the performance and clinical utility of OGM and the 523-gene NGS panel for the comprehensive genomic profiling of 15 myeloid cancers and compared it to our current diagnostic workflow that includes standard-of-care cytogenetic methods (karyotyping and FISH) and a 54-gene NGS panel. Finally, we demonstrate the visualization of OGM and NGS within the same bioinformatic platform to streamline analysis and reporting.

## Materials and Methods

### Sample selection

In this retrospective study, 15 well-characterized bone marrow aspirate (BMA) samples were received in our clinical laboratory for routine cytogenetic analysis (karyotype and FISH) and molecular analysis (54-gene myeloid panel), that had abnormal cytogenetic and/or molecular profiles were included in the study. Of these 15 samples, eight were MDS, six were AML, and one was MPN, namely essential thrombocythemia (ET). These 15 samples were processed for OGM and 523-gene NGS panel, with the technologist blinded to previous SOC results. The study was approved by the IRB A-BIOMEDICAL I (IRB REGISTRATION #00000150), Augusta University. HAC IRB # 611298. Based on the IRB approval, the need for consent was waived; all PHI was removed, and all data was anonymized before accessing for the study.

### Optical genome mapping

Ultra-high molecular weight (UHMW) DNA was isolated, labeled, and processed for analysis on the Bionano Genomics Saphyr® platform following the manufacturer’s protocols (Bionano Genomics, San Diego, USA). Briefly, a frozen BMA aliquot (650μl) was thawed and cells were counted using HemoCue (HemoCue Holding AB, Ängelholm, Sweden). Subsequently, a BMA aliquot comprising of approximately 1.5 million nucleated white blood cells was centrifuged, the cells were digested with Proteinase K, and lysed using LBB buffer. DNA was precipitated on a nanobind magnetic disk using isopropanol and washed using buffers (buffer A and B). The UHMW bound DNA was suspended in elution buffer and quantified using Qubit broad range (BR) dsDNA assay kits (ThermoFisher Scientific, San Francisco, USA).

DNA labeling was performed following manufacturer’s protocols (Bionano Genomics, USA) in which 750 ng of purified UHMW DNA was labeled at specific 6-base sequence motif with DL-green fluorophores using Direct Labeling Enzyme 1 (DLE-1) reactions. Following the labeling reaction, the DLE enzyme was digested using PK and the DL-green was removed in two steps using an adsorption membrane in a micro-titer plate. Finally, the DNA backbone was stained blue using DNA stain and quantified using Qubit high sensitivity (HS) dsDNA assay kits. Labeled DNA was loaded onto flow cells of Saphyr chips for optical imaging. The fluorescently labeled DNA molecules were imaged on the Saphyr platform after the labeled DNA molecules were electrophoretically linearized in the nanochannel arrays. Analytical QC targets were set to achieve >400X effective coverage of the genome, >70% mapping rate, 13-17 label density (labels per 100kbp), and >230 kbp N50 (of molecules >150 kbp).

### OGM variant calling and data analysis

Genome analyses was performed using Bionano Access (v.1.6)/Bionano Solve (v.3.6) software, and the rare variant analysis pipeline for all of the samples to assess and interrogate SVs and CNVs. Briefly, molecules of a given sample dataset were directly aligned to GRCH38, reference human genome assembly. SVs were identified where the pattern of labels in the molecules differed from the GRCH38 reference genome. Insertion, duplications, deletions, inversions, and translocations were called based on this alignment. SVs generated by the rare variant pipeline were then annotated with known canonical gene sets extracted from the reference genome assembly and compared to a control dataset to estimate the population frequency of SVs. Additionally, a coverage-based algorithm was used to call large CNVs (>5 kbp). A standard operating procedure (SOP) was devised to enable analysts to systematically and efficiently select for rare (<1% population frequency) variants with additional criteria that included filtering for variants overlapping genes.

### 523-gene NGS panel

DNA was isolated from BMA using the QIAamp DNA Blood Mini kit (QIAGEN, Hilden, Germany) as per the manufacturer’s protocol. Double-stranded DNA was measured using Qubit dsDNA broad-range assay kit (#Q32850, Invitrogen, USA) and 120 ng gDNA was used for library preparation. The libraries were prepared using the hybrid capture-based TSO 500 library preparation kit (# 20028214, TruSight Oncology 500 DNA Kit, Illumina, San Diego, CA) following the manufacturer’s instructions. In brief, the DNA was fragmented using an ultrasonicator (Covaris, Woburn, MA) with a target peak of ∼130 bp. After end repair, A-tailing, and adapter ligation, the adapter-ligated fragments were amplified using index PCR (UP-index) specific primers. Further, the libraries were enriched through a hybrid capture-based method using specific probes. This was followed by PCR-based enrichment, cleanup, and quantification of double-stranded DNA using high sensitivity Qubit (#Q32854 Invitrogen, USA) measurement. The libraries were subjected to bead-based normalization and were sequenced using V2 sequencing reagent kits on a NextSeq550 platform (Illumina, San Diego, CA) as per manufacturer recommendations.

### NGS variant calling and data analysis

The raw sequence reads FASTQ files were converted to BAM and VCF files using Qiagen clinical insight-interpret (QCI-I) clinical decision support software (Qiagen, Maryland, USA). The VCF files were analyzed using QCI-I for SNVs and small indels/duplications. Variants with a variant allele frequency (VAF) > 5% and a total read depth of >250X were filtered for analysis. The variants were categorized using evidence-based literature and manually curated data in QCI-I into tier classification based on AMP guidelines for classifying somatic variants. The variants were compared for concordance with previously reported variants, as the same samples were sequenced on a 54 gene myeloid panel and reported through PierianDx reporting solution.

### OGM and sequencing data: Visualization in NxClinical software

NxClinical v6.1 software is a unique data analysis and interpretation solution that can accommodate CNV and sequencing variants simultaneously from a single sample. Briefly, the variant call format (VCF) files from OGM and NGS data were uploaded into NxClinical and the data was analyzed for compound heterozygous events. The current iteration of NxClinical does not support the SV calls from OGM data; hence Bionano access (v6.1) was utilized for OGM data, QCI-I for sequencing data, and NxClinical for combined CNV and sequencing data viewing and analysis of compound heterozygous events overlapping CNV segments. In addition to simultaneous visualization of CNV and sequencing variants, BAM files from the NGS panel were utilized to call CNVs in NxClinical using the baseline sequencing reads throughout the genome. This approach was used to confirm the novel CNV calls detected by OGM with an alternative method (NGS).

## Results

### OGM quality control metrics and variant filtering

A typical run for OGM included the processing of eight samples in a batch for DNA isolation, and 12 samples for labeling, while three samples were loaded into one nanochannel chip onto the Saphyr instrument at a given time. All fifteen samples passed the quality control metrics with an average N50 (>150 kb) of 334 kb (±23), map rate of 89.6% (±5.8), label density of 16.9/100 kb (±1.1), and average coverage of 430x (±50). In total, 23030 SVs were identified in fifteen samples, with an average of ∼1520 SVs and ∼15 CNVs per sample. Of all the identified SVs, a total of 798 SVs and 224 CNVs survived the filtration criteria (96.4% variants filtered out), with an average of ∼53 SVs and ∼15 CNVs per sample that were further interrogated.

### OGM results: Concordance, higher resolution/resolving identified events, and additional findings

OGM achieved a 100% concordance with karyotyping and FISH in identifying all clinically reported SVs and CNVs in all fifteen cases (**Table 1**). The fifteen cases included ten simple cases (<3 aberrations) and five complex cases (>3 aberrations), based on aberrations detected with karyotype. OGM achieved 100% technical and clinical concordance in identifying all SVs that included interstitial deletion, duplication, translocations, and aneuploidies. In complex cases, OGM achieved 100% concordance in identifying all clinically relevant aberrations, while resolving previously observed aberrations with higher resolution. In a case of AML (case 7) with a complex karyotype of 45,XY,-5,-11,-17,add(18)(p11.3),-20,+3mar[19]/46,XY[1], with loss of 5q and gain of 5p by FISH, OGM identified gain of 5p, loss of 5q, complex rearrangements on chromosomes 11, 17, and 20 with several intra-and inter-chromosomal translocations (thus, resolving -11, -17, -20, and identifying the three marker chromosomes as 11, 17, and 20), and add(18)(p11.3) as an amplification at 18p11.3 with a CN state of 8 (**Figure 1 a-c**). In a case of MDS (case 9) with complex karyotype of 45,XX,-7[9]/44,XX,-5,del(7)(q11.2),-17,-18,+mar[3]/46,XX[8], with two abnormal clones, one with -7 and one with -5 and -7q by FISH, OGM identified loss of chromosome 7, and der(18)t(7;18)(q11.21;q12.2) resolving the other clone with del(7)(q11.2) and -18, and der(5;17) resolving -5, -17, and identifying the marker chromosome.

**Table 1.**
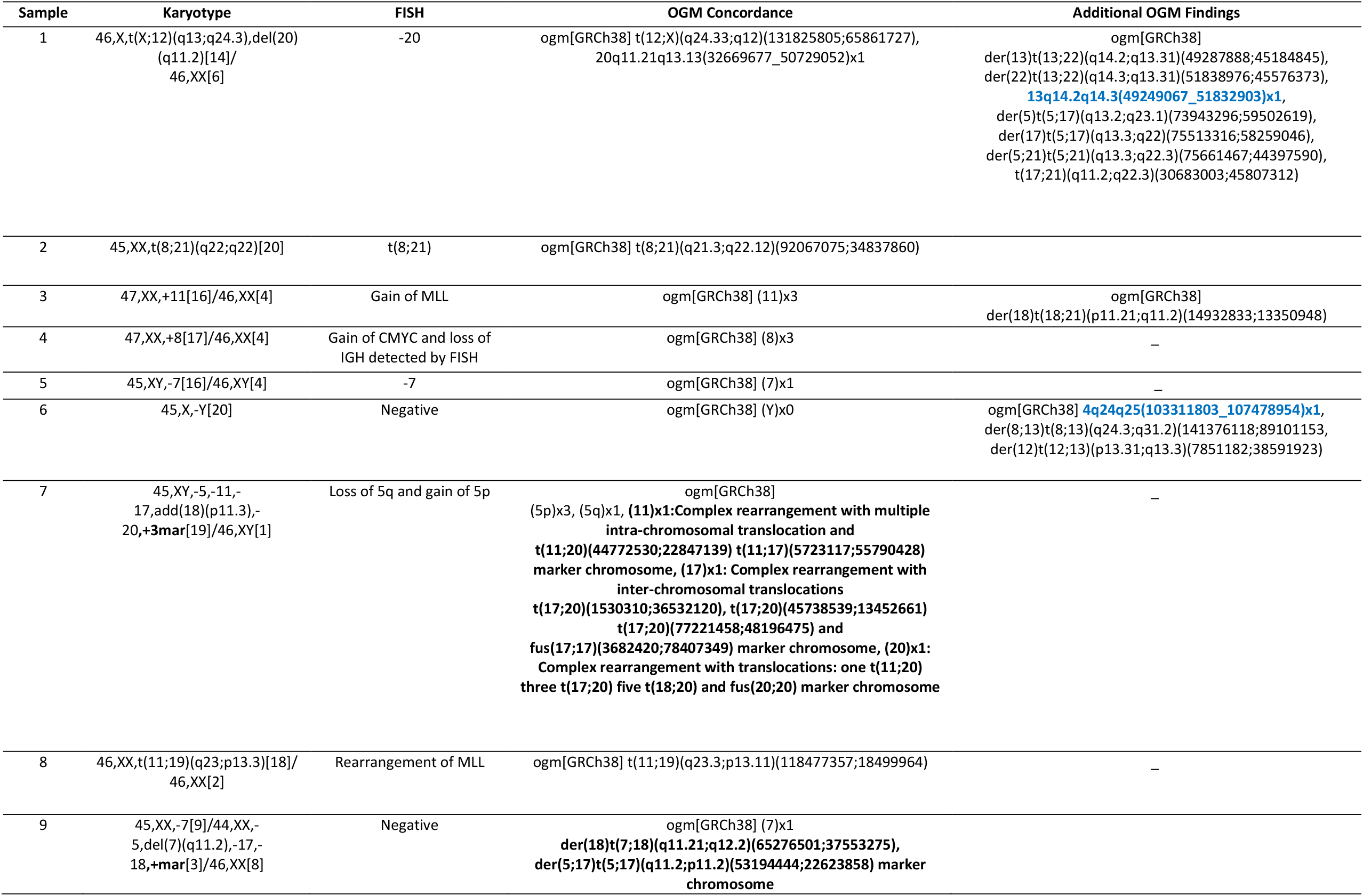
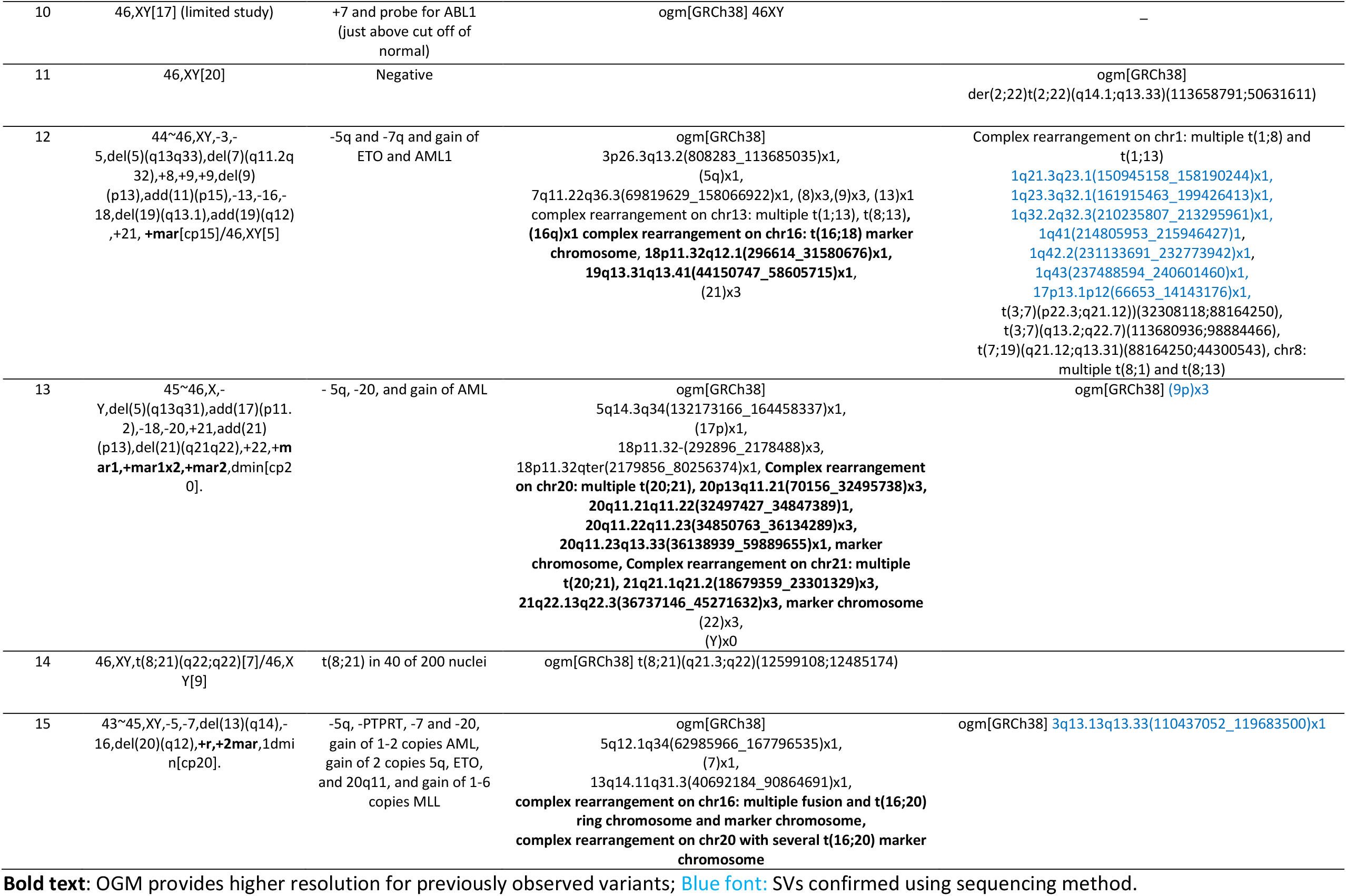
Comparison of optical genome mapping with Karyotype and FISH for cytogenetic analysis of myeloid cancers.

**Figure 1.**
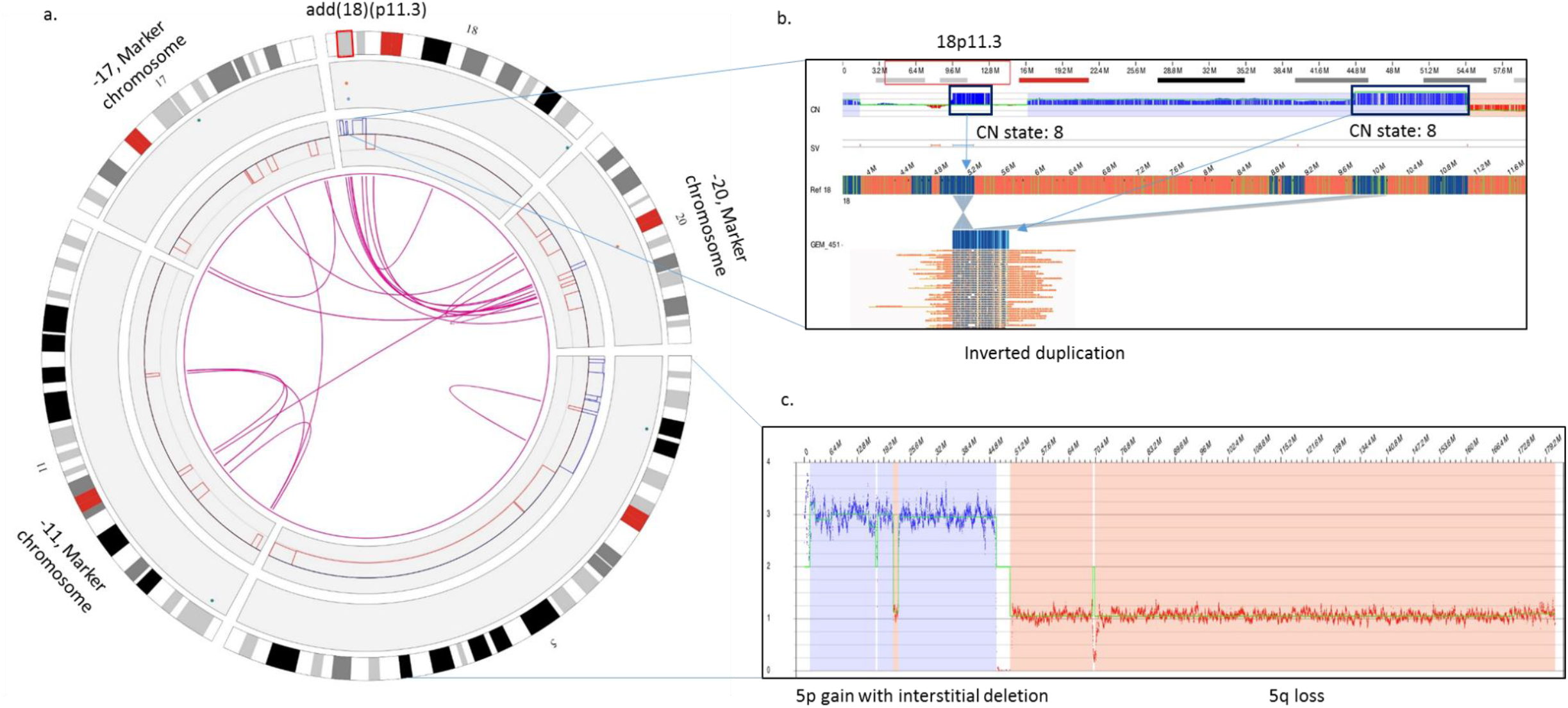
Optical genome mapping resolves a complex case of AML with higher resolution as compared to Karyotype and FISH. **a)** The circos plot summarizing the SVs identified in the genome: copy number gain on 5p, copy number loss on 5q, amplification on 18p, and complex rearrangements on chromosomes 11, 17, and 20. Optical genome mapping identifies the identity of marker chromosomes as 11, 17, and 20. **b)** zoomed-in view of 18p showing the amplification and complex rearrangement at 18p11.3 with a CN state of 8 of both the fused regions of the genome. **c)** Zoomed-in view of chromosome 5 showing copy number gain on 5p with an interstitial deletion and a copy number loss on 5q.

In addition to the previously identified SVs and CNVs, OGM identified several additional translocations and 11 copy number changes. Although the translocations have not been validated (beyond the scope of this study), all eleven additional CNVs were confirmed with the copy number signals from the CNV algorithm in NxClinical software using the sequencing data (**Figure 2 and 3**).

**Figure 2.**
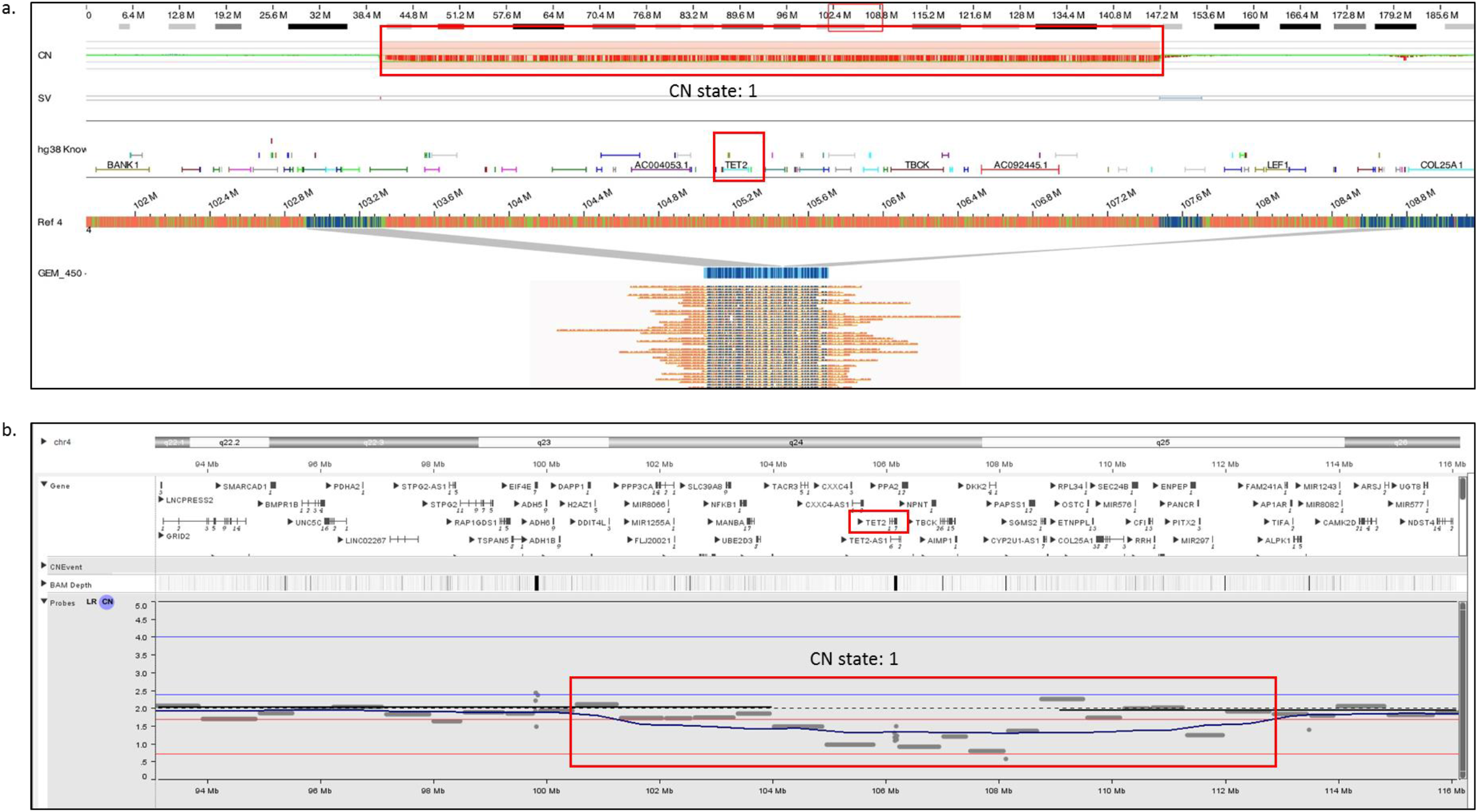
Optical genome mapping identifying a copy number loss on chr 4 [4q24q25(103311803_107478954)x1) deleting the *TET2* gene, which was missed by karyotyping. **b)** The CNV signals from the sequencing data visualized in the NxClinical software was indicative of the copy number loss and confirmed the deletion detected by optical genome mapping.

**Figure 3.**
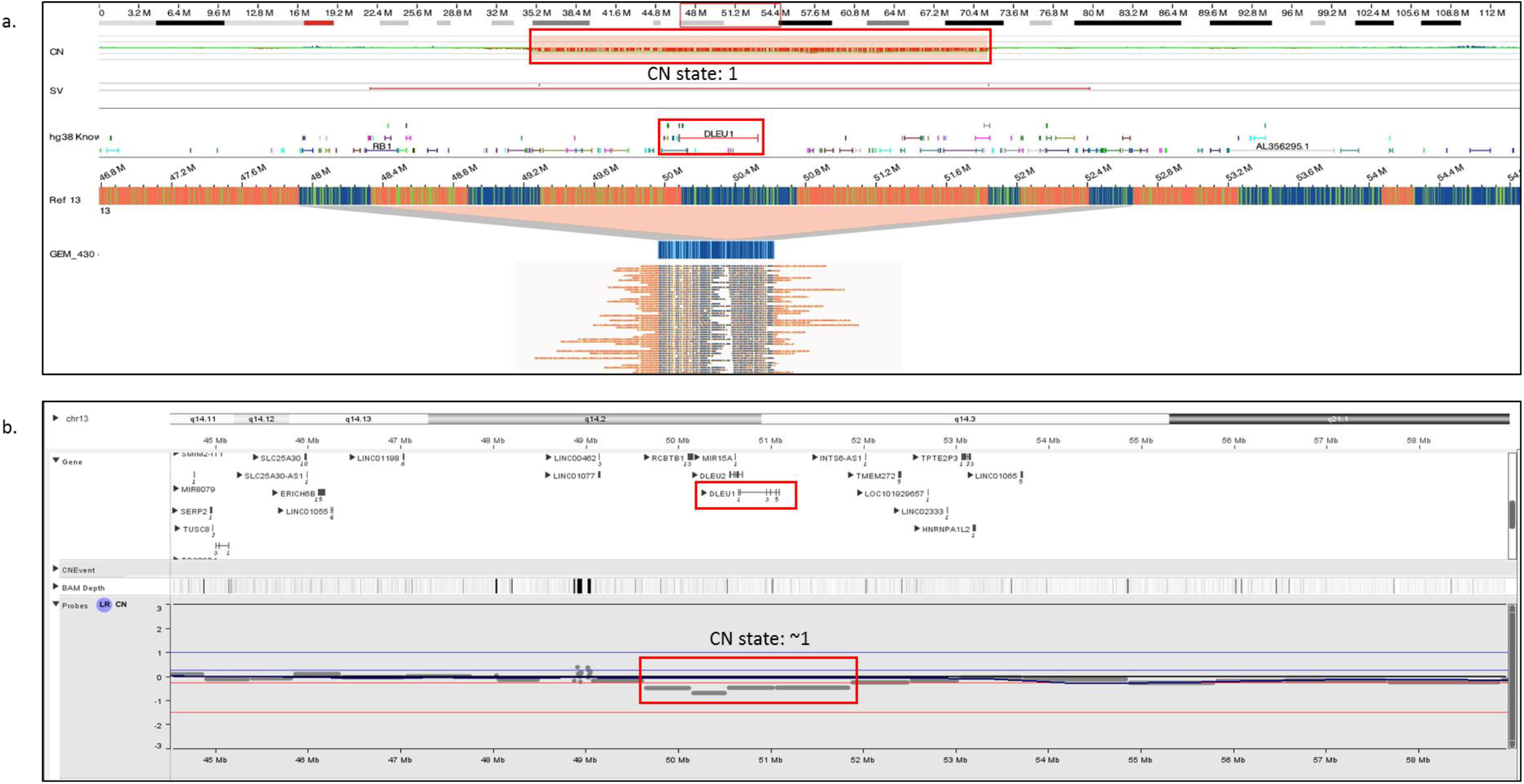
Optical genome mapping identifying a copy number loss on chr 13 [13q14.2q14.3(49249067_51832903)x1] deleting the *DLEU1* gene, which was missed by karyotyping. **b)** The CNV signals from the sequencing data visualized in the NxClinical software was indicative of the copy number loss and confirmed the deletion detected by optical genome mapping.

### 523-gene NGS panel quality control metrics

A typical sequencing run of the 523 gene NGS panel performed on the NextSeq550 platform consisted of 10 samples. The average percentage reads passing filter (PCT_PF) for a typical run was observed to be 90.1%. The percent base calls with a quality score of Q30 or higher for read 1 and 2 were 93.6 and 91.3, respectively. The four critical DNA library QC parameters viz. the median insert size from the sequencing reads for the run was found to be 125.3 bp; the average usable MSI counts were found to be 119.3; percent of exon bases with coverage >50X and percent target bases with coverage >250X were found to be 99.35 and 96.01, respectively. The variants were filtered in QCI-I using an built-in decision tree, filtering for rare somatic variants with a VAF ≥ 5%, read depth (>250x), and classified into tier-based classification, with the analyst and a board-certified director reviewing each call and classification.

### 523-gene NGS panel: Concordance and additional findings

The 523-gene NGS panel achieved 100% concordance in identifying the 22 previously identified clinically relevant sequence variants in these 15 samples (**Table 2**). QCI-I was 100% concordant in classifying the variants into correct tier-based classification compared to expertly classified reported variants. Further, the panel was able to identify seven additional clinically relevant variants (two tier 1A variants and five tier 2C variants) in four samples. 2/7 of these variants were in genes that were not covered in the 54-gene panel.

**Table 2.** Comparison of 523-gene NGS panel with 54-gene NGS panel for detecting clinically relevant variants in myeloid cancers.

| Sample | Phenotype | 54 gene NGS panel (reported variants) | 523-gene NGS panel Concordance | Tier classification | 523-gene NGS panel additional variants | VAF (%) | Tier classification* |
| --- | --- | --- | --- | --- | --- | --- | --- |
| 1 | MDS | MPLp.W515L | MPLp.W515L | 1A | ASXL1p.Q428fs*10 | 29 | 1A |
|  |  |  |  |  | PMS2p.L729fs*6 | 16 | 2C |
|  |  |  |  |  | SH2B3p.C133fs*64 | 13 | 2C |
| 2 | AML | — | — |  | RAD21p.E99fs*3 | 26 | 2C |
| 3 | AML | NRASp.Q61R | NRASp.Q61R | 2C |  |  |  |
|  |  | NRASp.G12D | NRASp.G12D | 2C |  |  |  |
|  |  | IDH2p.R172K | IDH2p.R172K | 1A |  |  |  |
| 4 | AML | JAK2p.V617F | JAK2p.V617F | 2C |  |  |  |
|  |  | IDH2p.R172K | IDH2p.R172K | 1A |  |  |  |
| 5 | AML | DNMT3Ap.R882H | DNMT3Ap.R882H | 1A |  |  |  |
|  |  | IDH2p.R140Q | IDH2p.R140Q | 1A |  |  |  |
|  |  | NPM1p.W288fs*? | NPM1p.W288fs*? | 1A |  |  |  |
|  |  | NRASp.G12V | NRASp.G12V | 2C |  |  |  |
|  |  | PTPN11p.F71L | PTPN11p.F71L | 2C |  |  |  |
| 6 | MDS | KRASp.A146V | KRASp.A146V | 2C |  |  |  |
|  |  | KRASp.K117N | KRASp.K117N | 2C |  |  |  |
|  |  | TET2p.S1132Lfs*6 | TET2p.S1132Lfs*6 | 2C |  |  |  |
| 7 | AML | NRASp.G13D | NRASp.G13D | 2C | TP53c.97-1G>T<br>KRASp.G13D | 89<br>25 | 1A<br>2C |
| 8 | AML | TET2p.N535Kfs*7 | TET2p.N535Kfs*7 | 2C |  |  |  |
| 9 | MDS | TP53p.V173L | TP53p.V173L | 1A |  |  |  |
| 10 | ET | CALRp.K385Nfs | CALRc.1154_1155insTTGTCp.K385fs*? |  |  |  |  |
| 11 | MDS | — | — |  |  |  |  |
| 12 | MDS | TP53c.646G>A<br>p.V216M | TP53c.646G>A<br>p.V216M | 1A |  |  |  |
| 13 | MDS | — | — |  |  |  |  |
| 14 | MDS | FLT3-TKD pD835Y | FLT3c.2503G>T<br>p.D835Y | 1A | RAD21c.764T>A<br>p.L255* | 23 | 2C |
| 15 | MDS | TP53p.A159V | TP53c.476C>Tp.A159V | 1A |  |  |  |
|  |  | TP53p.D281V | TP53c.842A>Tp.D281V | 1A |  |  |  |

### OGM and 523-gene NGS panel: Simultaneous visualization and CNV confirmation in NxClinical

The simultaneous visualization of CNVs and sequencing variants (SNV and small indels) from OGM and 523-gene NGS panel, respectively in the NxClinical software identified two clinically relevant compound heterozygous events in two samples. In a case of MDS (sample 6), OGM detected a heterozygous interstitial deletion on chr 4 [4q24q25(103311803_107478954)x1], which included the *TET2* gene, while the sequencing data detected a pathogenic frameshift variant (*TET2*p.S1132Lfs*6, Tier 2C classification) on the other allele (**Figure 4a**). Further, the deletion on chr 4 detected by OGM was also confirmed using the sequencing reads (**Figure 2a-b**). It must be noted that this cryptic deletion cannot be identified by karyotyping and is not part of the targeted FISH panel. In the case of AML (sample 7), OGM identified a complex rearrangement on chr 17 involving the deletion of the *TP53* gene, while NGS detected a pathogenic splice site alteration (*TP53*c.97-1G>T; Tier 1A classification) [**Figure 4b**].

**Figure 4.**
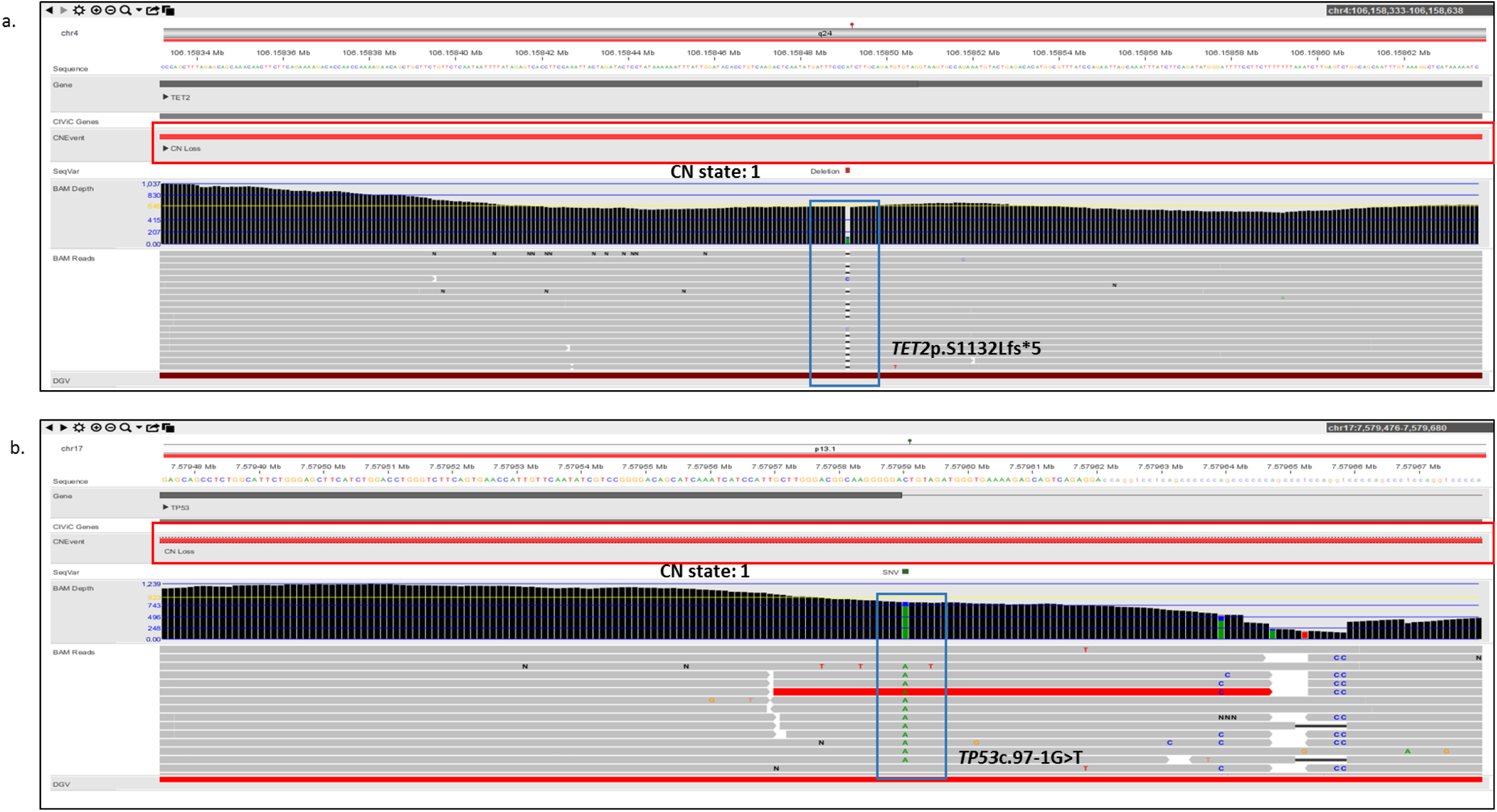
The combined visualization of copy number changes and single nucleotide variants in NxClinical identified compound heterozygous events in two samples. **a)** The copy number loss is highlighted in red, with a CN state of 1, while the *TET2*p.S1132Lfs*5 variant is highlighted in blue. **b)** The copy number loss is highlighted in red, with a CN state of 1, while the *TP53*c.97-1G>T splice variant is highlighted in blue.

## Discussion

The present study aimed at evaluating a cost-effective and high yield approach of combing OGM and 523-gene NGS panel as an alternate diagnostic workflow compared to the current standard of care cytogenetic and molecular technologies for genomic characterization of myeloid cancers. This study demonstrated the following: a) higher resolution of OGM in detecting SVs and CNVs as compared to the combination of current standard-of-care cytogenetic techniques (karyotyping and FISH), b) greater clinical utility of the 523-gene NGS panel as compared to a 54-gene NGS panel for detecting clinically relevant sequencing variants, and c) greater clinical utility of combining OGM with a 523-gene NGS panel for a comprehensive approach to a higher success rate in the profiling of myeloid cancers.

### Cytogenomics: OGM compared to standard-of-care technologies (karyotype and FISH)

The sample processing for OGM is simple and standardized for BMA and blood samples and can be easily employed by any diagnostic molecular or cytogenetic laboratory for clinical implementation. Typically, 16 samples were isolated in a single day in two batches of 8 samples, each, and 12 samples were efficiently labelled in a single batch. Overall, 12 samples were labelled over 2 days, with a total hands-on time of approximately 5 hours. Three samples were loaded on a single Saphyr chip and imaged on the instrument in a single run, with ∼1500 gigabases of data collected for each sample enabling an average depth of coverage of ∼430x. All 15 samples passed the pre-analytical and analytical QC metrics, which highlights the ease of use and demonstrates the feasibility of processing blood and BMA samples for cytogenetic analysis in a routine laboratory. The entire OGM workflow from sample to SV calls can be accomplished in 4 days.

OGM achieved 100% concordance in detecting all clinically relevant SVs and CNVs in the cases with simple cytogenetic profiles while demonstrating a higher resolution in resolving the genetic aberration in cases with the complex cytogenetic profile. These results are in agreement with recently published work on the characterization of hematological malignancies [**15-17**]. The cytogenetic profile obtained with karyotyping and FISH in complex cases could not identify or detect the cryptic, clinically relevant aberrations, while it was only after the analysis on OGM that the “true” complex nature of these genetic aberrations was revealed. As recently demonstrated, we found that OGM could discern complex rearrangements and resolve previously identified genetic aberrations with much higher resolution. OGM could resolve the identity of marker chromosomes in all five complex cases. Notably, we would like to highlight that in a complex case of MDS, OGM was able to resolve the karyotype and reveal the identity of the marker chromosome that was detected in 3/20 cells (at 7.5% allele fraction) by karyotyping. Although the limited number of metaphase cells analyzed in a karyotype analysis is insufficient to ascertain the true level of mosaicism, OGM performed at ∼400x depth assisted in the discovery of the content of the marker chromosome at an apparent 7.5% allele fraction.

Overall, OGM offers several advantages over karyotyping and FISH for genomic characterization of myeloid cancers. First, OGM does not require cultured cells for genomic characterization, as required with karyotyping and FISH. The elimination of this pre-analytical step leads to reduced turnaround time compared with SOC methods. Second, OGM demonstrates the unique ability for genome-wide analysis of all classes of SVs and CNVs at high resolution in a single assay. Third, OGM demonstrates higher resolution in resolving complex genetic aberrations and reveals these events with greater accuracy compared to karyotype. Fourth, OGM demonstrates the ability to detect both balanced and unbalanced translocations in a single assay with precise breakpoints, sufficient for the detection of novel gene fusions. OGM can also discern the location and orientation of copy number gains (distinguishing tandem and inverted duplications that might have different functional implications). Based on our experience, OGM is a relatively simple technique compared to karyotype and FISH and requires minimal laboratory setup and technical experience to perform the assay.

### Molecular profiling: 523-gene NGS panel compared to 54-gene NGS panel

The comprehensive molecular profiling of myeloid cancers is of significant interest and owing to the decreasing cost of sequencing technologies, it is now possible to evaluate and implement comprehensive gene panels, exome sequencing (ES), or genome sequencing (GS) for clinical care. However, because of data analysis and reporting complexities, limited gene (≤54 genes) panels have been routinely employed by clinical laboratories. As we have previously demonstrated [**7**], the 523-gene NGS panel achieved excellent quality control metrics and demonstrated a 100% concordance in identifying clinically relevant variants previously identified with the 54-gene panel. In addition, we demonstrate the ability of a larger gene panel in detecting additional clinically relevant sequencing variants as compared to panels that are limited in gene content. The 523-gene panel, not only has a higher gene content, but also has a higher genic coverage compared to the 54-gene panel, which enables the detection of clinically relevant SNVs in known hotspot genes.

Further, we demonstrate the use of QCI-I as a data analysis and reporting solution that demonstrated a 100% concordance in classifying variants into correct tier classification as compared to expert classification, as demonstrated previously. Overall, the panel has extensive coverage across the entire genome, for variants significantly beyond those captured on existing NGS platforms for hematological malignancies. The 523-gene panel has several advantages as compared to the 54-gene panel that includes: a) the ability to detect additional clinically relevant variants in genes currently not covered in smaller gene panels and b) the 523-gene panel is a comprehensive pan-cancer panel that includes genes implicated across cancers and has the potential to identify variants that would potentially affect diagnostic, prognostic, and/or therapeutic course for management of these malignancies. Molecular genetic or pathology laboratories are already equipped with tools for sequencing small gene panels and are familiar with library preparation and sequencing, hence the adoption of a larger gene panel would not lead to any major changes to the lab workflow.

### OGM and 523-gene NGS panel compared to the current diagnostic workflow (karyotype, FISH, and 54-gene NGS panel)

The combination of OGM and the 523-gene NGS panel demonstrates several advantages as compared to current standard-of-care technologies. Apart from the higher resolution and comprehensive genome-wide coverage of SVs via OGM, the simultaneous use of these technologies quickly identified compound heterozygous events that were not interrogated with routine methods. *TET2* is an important gene implicated in MDS and is routinely included in the NGS panels for detecting sequencing variants. In one case, OGM uniquely identified a large deletion of 4.1 Mb (missed by karyotyping and not interrogated by FISH) and NGS identified a LoF variant (detected by both small and large panels). By visualization in the NxClinical platform, observation of both variants allowed easy determination of compound heterozygosity involving this gene in the tumor (Figure 4a). In an AML case, however, loss of chromosome 17 (indicative of loss of *TP53*) was detected by karyotyping, which was resolved as a complex rearrangement of chr 17 with the loss of one copy of *TP53* gene by OGM. The 523 gene NGS panel identified a splice variant in this gene that was missed by the 54-gene NGS panel. By visualization in the NxClinical platform, observation of both variants allowed easy determination of compound heterozygosity involving this gene in the tumor (Figure 4b). Although, the study is limited in the number of cases evaluated, resolving the genomic complexity in these two cases (using both OGM and the large NGS panel) highlights the benefit of obtaining clinically relevant information, on top of their respective advantages. Further, the additional CNVs detected by OGM were confirmed with the copy number signals in NxClinical using the sequencing data. Notably, the 523-gene NGS panel does not have a genome-wide coverage and thus, the sequencing data could only be indicative of copy number changes that can be used to confirm the calls originally detected by OGM. Overall, these two technologies complement each other and enable comprehensive and robust profiling of myeloid cancers in clinical care (**Figure 5**).

**Figure 5.**
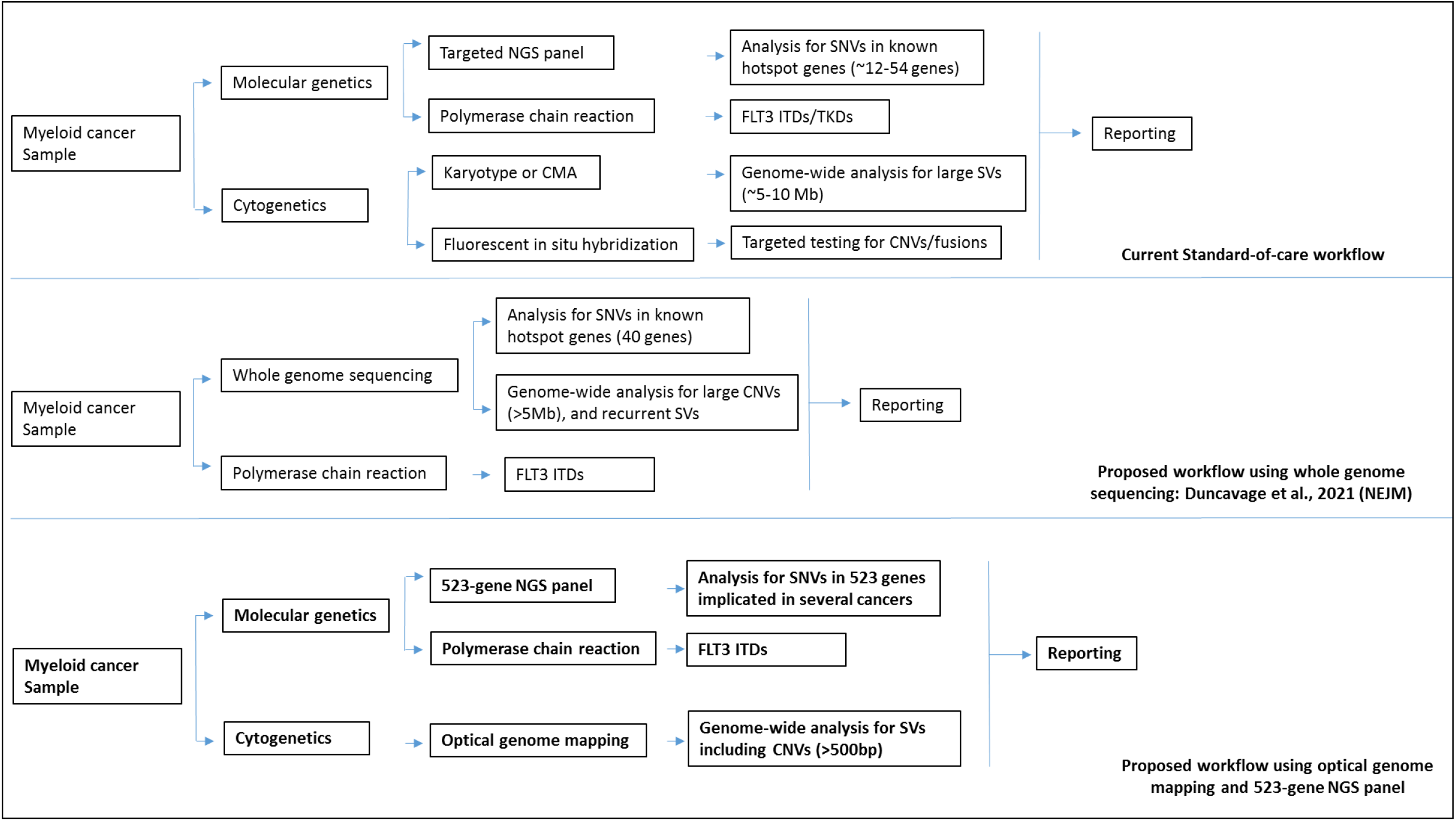
Schematic depicting the various workflows for the genetic testing of myeloid cancer samples. Top panel: current standard-of-care workflow. Middle panel: workflow proposed by Duncavage et al., 2021 using whole-genome sequencing. Bottom panel: proposed workflow with optical genome mapping and 523-gene NGS panel.

### OGM and 523-gene NGS panel compared to GS for myeloid cancers

A recent study demonstrates the use of GS to detect both cytogenetic (karyotype level) and sequencing variants in a single assay in myeloid cancers, as compared to karyotyping and a 40-gene NGS panel [**18**]. The results of GS were encouraging but several challenges need careful consideration before implementing for clinical use. Herein, we compare OGM and 523-gene NGS panels to GS for genomic profiling of myeloid cancers. Comparison of GS with MyeloSeq (40 gene NGS panel), GS at ∼50x depth failed to detect several actionable sequencing variants across a wide range of VAF(false-negative rate). Upon increasing GS coverage depth (∼120x), better performance was achieved but an important *KRAS* variant (detected with Myeloseq at 5.2% VAF) was still missed. The failure was attributed to the lower depth of coveregae of GS (∼120x vs 500x), compared with MyeloSeq. We suggest that the 523-gene panel seems to be an ideal alternative that achieves coverage of >250x across the targeted genes and provides a comprehensive assessment of sequencing variants without compromising clinically relevant variants present at low VAF representing the heterogeneous tumor burden in these malignancies. For cytogenetic characterization, GS at ∼50x depth was concordant in detecting CNVs and translocations detected with karyotype at the 5 Mb resolution (LoD for karyotype). However, the data analyzed by GS was limited to interrogating recurrent SVs only. Non-recurrent SVs, CNVs smaller than 5Mb, and cases with complex rearrangements/ marker chromosomes were not investigated or reported by GS. This limited comparison of WGS performance with >5Mb abnormalities in karyotype is a fundamental flaw in the study and approach for investigating myeloid cancers. Given that ∼45% of myeloid cancers remain cytogenetically normal, GS (at the current depth of coverage and exhaustive bioinformatics capabilities needed for SV detection) does not offer additional benefits over the traditional cytogenetic methods. Multiple studies have demonstrated that OGM can detect clinically actionable SVs and novel fusions that can affect the risk stratification and management of these cancers. Overall, the combined cost for OGM and the 523-gene NGS panel is lower than GS at 50x coverage (requiring a sophisticated bioinformatics core at every laboratory) and offers several advantages w.r.t detecting clinically relevant variants, data analysis, and reporting (**Table 3, Figure 5**). Based on current data and expectation from other studies, we expect that GS at 50X or 120X depth will continue to miss some sequence variants detected by high depth gene panel as well as structural variations that are intractable to GS, while OGM + 523 gene panel will provide all SOC variants as well as additional important clinically relevant variants.

**Table 3.** Comparison of the proposed workflow of optical genome mapping and 523-gene NGS panel with current diagnostic workflow and workflow proposed by Duncavage et al., 2021 using whole-genome sequencing.

| Variant Classification | Variant types | Variant Classes | Current diagnostic workflow |  |  | Duncavage et al., 2021 | Proposed workflow |  |
| --- | --- | --- | --- | --- | --- | --- | --- | --- |
|  |  |  | Karyotype | FISH | 54-gene NGS panel | short-read WGS | OGM | 523-gene NGS panel |
| Sequencing variants | SNVs | – | – | – | 500x cutoff | 50x (probability of false negatives for variants detected at low VAF with 500x panels) | – | 250x cutoff |
|  | Indels | – | – | – | – | – | – | – |
| Aneuploidy | Monosomy | Monosomy | √ | Targeted FISH probes | – | √ | √ | – |
|  | Trisomy | Trisomy | √ | Targeted FISH probes | – | √ | √ | – |
|  | Triploidy | Triploidy | √ | Targeted FISH probes | – | √ | no | – |
|  | Tetraploidy | Tetraploidy | √ | Targeted FISH probes | – | √ | no | – |
|  | Ring chromosome | Ring chromosome | √ | No | – | No | ≥500 kbp + fusion break | – |
| Copy Number Variants | Deletions/Duplication | Interstitial | 5Mb or larger | Targeted FISH probes | – | 5Mb or larger | ≥500 bp | – |
|  |  | Terminal | 5Mb or larger | Targeted FISH probes | – | 5Mb or larger | ≥500 bp | – |
|  | Insertion | Interstitial (unknown sequence) | 5Mb or larger | No | – | No | ≥500 bp | – |
|  |  |  |  |  | – |  |  | – |
| Structural variants | Translocations | Balanced translocations | √ | dependent on FISH probes | – | Only recurrent translocations have been investigated with current bioinformatics processing | √ | – |
|  |  | Unbalanced translocations | √ | dependent on FISH probes | – | Only recurrent translocations have been investigated with current bioinformatics processing | √ | – |
|  | Inversions | Pericentric |  |  | – | No | √ | – |
|  |  | Paracentric | 5Mb or larger | No | – | No | ≥30 kbp | – |
| Homozygosity mapping | LOH | AOH/ROH/LOH | No | No | – | √ | Not currently | – |
| Microsatellite/Macrotaellite repeats | Repeats | Expansions/Contractions | No | No | – | Limited to short repeats | ≥500 bp | – |

Our study has a few limitations, first, the study is performed on a limited number of cases and a validation of this workflow with a higher number of samples would be needed. We have performed a literature review and compared the internal data from the proposed combination workflow of OGM plus a 523-gene NGS panel to GS; however, an ideal comparison would be to perform both workflows on the same samples. The additional translocation (novel fusion event) identified in this study needs to be validated and the functional implications of novel gene fusions need to be investigated. Even though this was beyond the scope of our study, we have shared all relevant information for future discovery. Overall, this proof-of-principle study demonstrating the use of OGM and 523-gene NGS panel for comprehensive genomic profiling of myeloid neoplasm shows immense potential to be integrated into a routine cytogenetic laboratory for maximum impact of disease diagnosis and management.

## Data Availability

All relevant data has been included in the manuscript.

## Ethics approval

The study was approved by the IRB A-BIOMEDICAL I (IRB REGISTRATION #00000150), Augusta University. HAC IRB # 611298. Based on the IRB approval, the need for consent was waived; all PHI was removed, and all data was anonymized before accessing for the study.

## Declarations

RK has received honoraria, and/or travel funding, and/or research support from Illumina, Asuragen, QIAGEN, Perkin Elmer Inc, Bionano Genomics, Agena, Agendia, PGDx, Thermo Fisher Scientific, Cepheid, and BMS. DS, SS, AH, and AC are salaried employee at Bionano Genomics Inc. All other authors have no competing interests to disclose.

